# Honey Experiment on LeProsy Ulcer (HELP) Study Protocol: A Randomised Control Trial of Raw, Unadulterated African Honey for Ulcer Healing in Leprosy: A Study Protocol

**DOI:** 10.1101/2023.07.14.23292603

**Authors:** Sunday Odihiri Udo, Pius Ogbu Sunday, Paul Alumbugu Tsaku, Israel Olaoluwa Oladejo, Anthony Meka, Linda Ugwu, Motunrayo Ajisola, Joshua Akinyemi, Akinyinka Omigbodun, Sopna Choudhury, Jo Sartori, Onaedo Ilozumba, Samuel I. Watson, Richard J Lilford

**Affiliations:** The Leprosy Mission Nigeria, Abuja, Nigeria; German Leprosy and Tuberculosis Relief Association/RedAid Nigeria, Enugu, Nigeria; Clinical Trials Unit, University of Ibadan, Oyo State, Nigeria; Institute of Applied Health Research, College of Medical and Dental Sciences, University of Birmingham, Birmingham, UK

**Keywords:** Honey, Leprosy, *Mycobacterium leprae*, Ulcer, Nigeria

## Abstract

**Background:** As in diabetes, ulcers in leprosy occur as a result of nerve damage and loss of sensation due to neuropathy. The aim of this study is to evaluate the healing properties of raw, undiluted African honey in comparison with normal saline dressing of leprosy ulcers.

**Methods:** This is a multi-centre, comparative, prospective, single-blinded, parallel-group, 1:1 individually randomised controlled trial to be conducted at The Leprosy Referral Hospital, Chanchaga, Niger state in central Nigeria and St. Benedict Hospital, Ogoja, Cross River state, South-south Nigeria. Raw, unadulterated honey will be used in the ulcer dressing of the eligible consenting participants in the intervention group while those in the control group will be treated by dressing with normal saline. The main outcomes will be the proportion of complete healing and the rate of healing up to 84 days post-randomisation. Follow-up will be for 6 months from randomisation. We aim to enter 90-130 participants into the study. Observations will be made by blinded observers examining photographs of ulcers.

**Discussion:** Our study will provide an unbiased estimate of the effect of honey on the healing of neuropathic ulcers.

**Trial registration:** ISRCTN10093277. Registered on 22 December 2021.

## Introduction

### Background and rationale {6a}

Leprosy or Hansen’s disease is a chronic granulomatous disease caused by *Mycobacterium leprae* [1]. The disease morbidity is characterised by damage to the skin and peripheral nerves, which results in neuropathy, severe disability, and the consequent social exclusion and stigmatization [2]. Leprosy is preventable and treatable with multidrug therapy (MDT) but remains endemic in many communities of people living in poverty and poor hygiene [2]. Nigeria reported a total of 1,417 new cases of leprosy in 2020, including 87 new child cases [3], with grade 2 disability rate at about 15% for the past 5 consecutive years [4].

Thirty – fifty percent of people infected with leprosy are reported to eventually have nerve damage or neuropathy [5]. Ulcerations usually occur in anaesthetic foot. Ulcers heal slowly with routine therapy and have a tendency to recur [6].

The use of honey as a therapeutic agent in the treatment of wounds dates back to ancient times with the earliest documented report recorded in Edwin Smith papyrus (2600 – 2200 BCE) [7]. Honey is a viscous, supersaturated solution containing sugars, water, amino acids, vitamins, minerals, enzymes, and many other substances which are derived from nectar gathered and modified by honeybee, *Apis mellifera* [8]; [9]. Studies have suggested that honey promotes wound healing, stimulates tissue growth, facilitates debridement and epithelisation, deodourises, reduces oedema and exudates, and possess antimicrobial properties [7;10;11].

There has been a recent resurgence of interest in the use of honey for treatment of different kinds of wound as researchers continue to search for improved, cost-effective, and readily available agents to promote wound healing [9;12]. Although there is a sizeable number of reports that shows mixed levels of effectiveness in the use of honey as a topical agent for treatment of different types of ulcers, our search of numerous databases revealed a paucity of documentation on the use of honey in the treatment of ulcers in leprosy. The effectiveness of honey to promote healing of ulcers in general remains uncertain. A Cochrane review has reported several studies with unclear outcomes for trials with honey on venous leg ulcers, diabetic foot ulcers, and mixed chronic wounds. Most of the reported studies were adjudged by the reviewers to be of low quality due to imprecision, and high risk of bias. The review suggested high risk of bias in the reports due to non-blinding of study participants and healthcare professionals. Statistical heterogeneity was evident across studies [9].

The current practice in the dressing of leprosy ulcers in Nigeria is the use of normal saline. Honey is acknowledged to be safe for use in wound dressings [15], with only about 5 % of patients having painful sensation after dressing, and undocumented concern of botulism disease due to infection *Clostridium botulinum* [16]. This study will evaluate the effectiveness of honey in treatment of ulcers in leprosy in comparison with normal saline dressing.

## Objectives {7}

The aim of this study is to evaluate the healing properties of raw, unadulterated African honey in comparison with normal saline dressing of leprosy ulcers.

The study objectives are:

i. Recruit 90 – 130 eligible, consenting people within 12 months.
ii. Randomise the participants to receive ulcer dressing with either honey (intervention group) or normal saline (control group) twice a week.
iii. Observe the rate of healing based on two observations per week (cm^2^ per unit time/until either the ulcer has healed or 84 days have elapsed since randomisation).
iv. Observe the time to healing defined as complete re-epithelisation (up to a maximum of 84 days).
v. Monitor the rate of activity of study participants with pedometers.
vi. Monitor recurrence of treated ulcers or appearance of new ulcers and any anatomical changes in the limb at 6 months follow-up after randomisation.

### Trial design {8}

The study is a multi-centre, prospective, single-‘blinded’, parallel group, 1:1 individually randomised controlled trial. Study duration is 48 months (maximum) with recruitment starting at month 5 and post-discharge follow-up starting at month 11. We aim to enroll between 90 and 130 participants over a 24 month recruitment period. The study pathway is shown in Fig. 1.

**Fig. 1:**
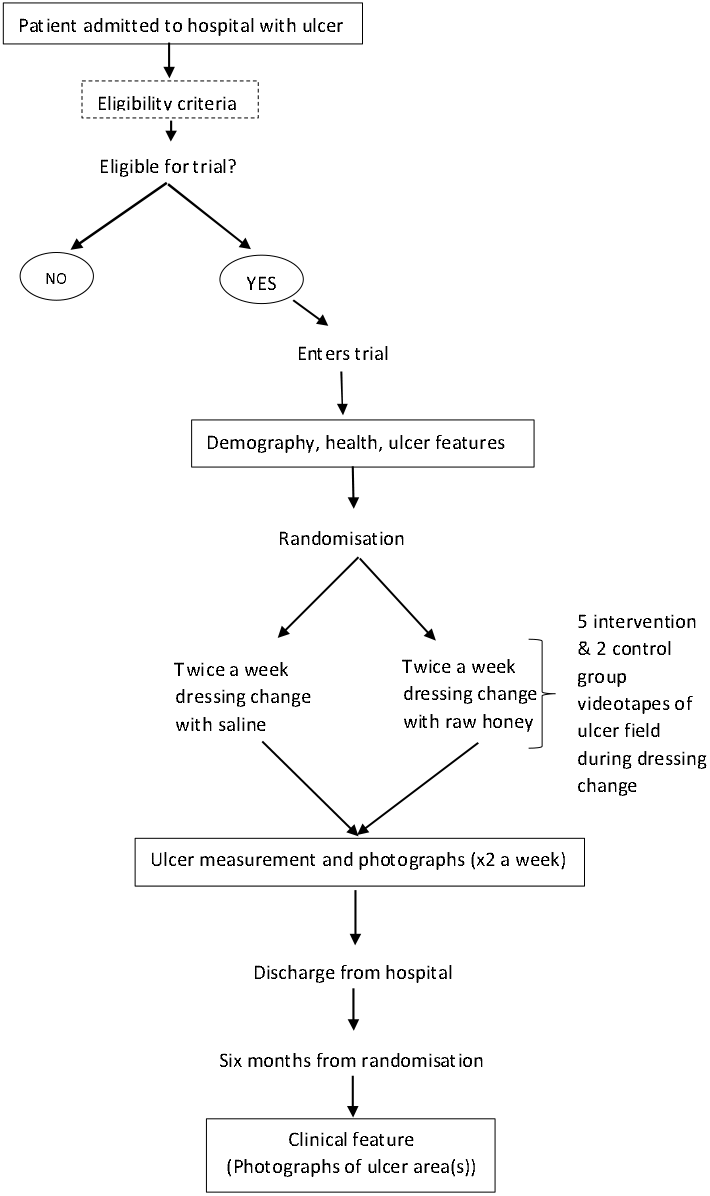
HELP study pathway

**Figure 2.**
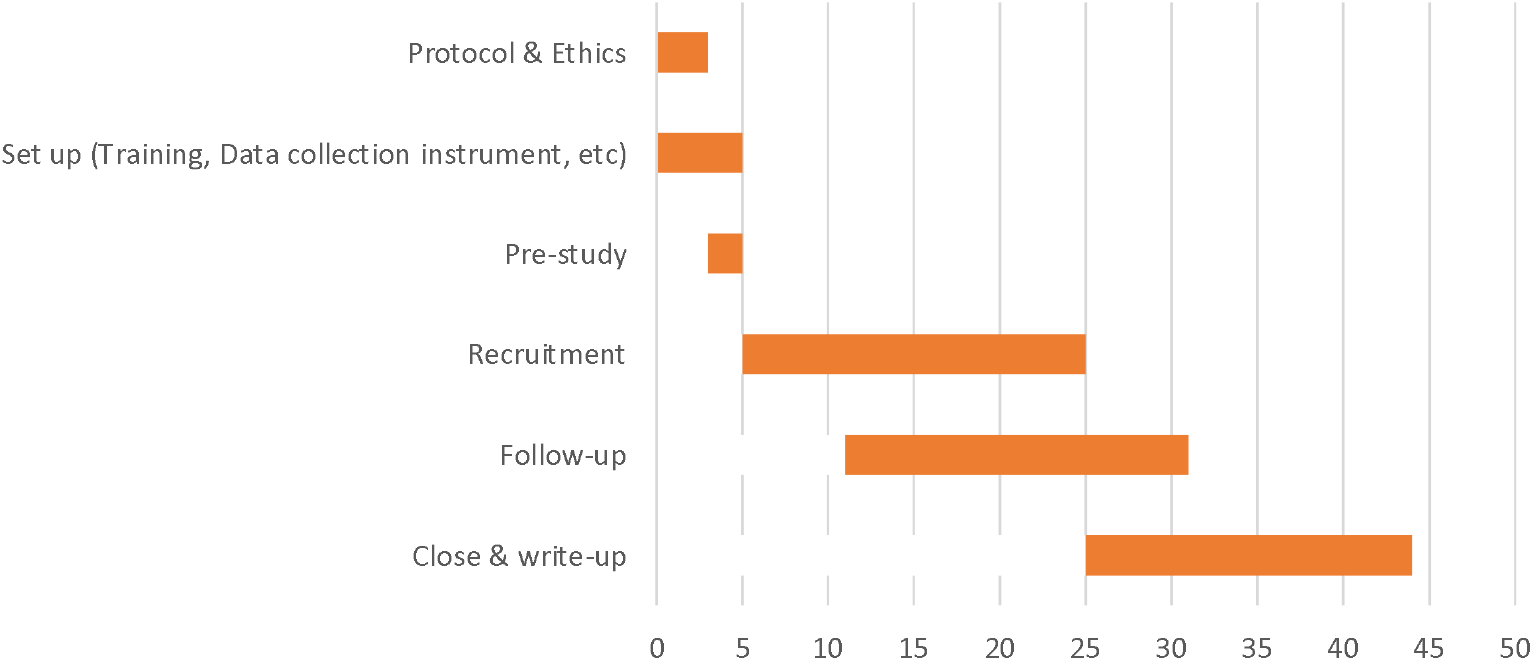
Timeline (in months) for HELP trial (based on most pessimistic estimate)

## Methods: Participants, interventions and outcomes

### Study setting {9}

The study centres are, The Leprosy Rehabilitation Hospital, Chanchaga, Minna, Niger state and St Benedit’s TBL and Rehabilitation Hospital, Ogoja, Cross River state. The main study will be preceded by pre-study diligence.

The Chanchanga Leprosy Hospital in Minna, Nigeria is a specialist hospital operated by the government of Niger state in collaboration with The Leprosy Mission Nigeria. The hospital was established in 1940 and has two wards, eye and physiotherapy units, theater, laboratory, and dispensary. The hospital is supported by an orthopaedic workshop which is built and managed by The Leprosy Mission Nigeria, which produces assistive devices including wheelchairs, crutches, protective sandals and artificial legs.

A doctor, physiotherapists, and nurses manage leprosy patients in the hospital, including those with severe ulcers. The regular practice for patients with chronic ulcers is soaking of the wounds, cleaning, and dressing with normal saline. Honey is readily available and has been used in some instances, but the usage did not follow any standardized guide and the outcomes were not documented.

St Benedict’s TBL and Rehabilitation Hospital is a TB and leprosy referral hospital located in Ogoja, a town in the northern part of Cross River State of Nigeria. It is owned by the Catholic Diocese of Ogoja, Cross River State, Nigeria. It provides these services in collaboration with the National Tuberculosis and Leprosy Control Programme (NTBLCP) and with the technical assistance of German Leprosy and TB Relief Association (DAHW). The centre has a community outreach programme which covers 31 communities in Bekwarra, Ogoja and Yala Local Government Areas of Cross River State. The facility is staffed by 2 medical doctors, 9 registered nurses, 5 trained community health extension workers (CHEWs), 3 auxiliary nurses, 5 ward orderlies, 1 lab technician, 2 lab attendants, 1 physiotherapy assistant, 1 shoe maker, 1 SER officer. It has a total of 97 beds, comprising 2 main wards of 25 beds each, 2 minor wards with 15 beds and a TB camp with 32 beds. It also has a surgical theatre, a physiotherapy section, laboratory, OPD and shoe making workshop.

### Eligibility criteria {10}

The local medically qualified researchers employed on the grant will screen all ulcer admissions. The researchers will complete online eligibility forms in respect of each patient with an ulcer. Consent will be sought by the research fellow from consecutive patients ≥18 years with a foot / leg ulcer of between 2 and 20cm2 and not requiring surgery (e.g. skin graft). The inclusion criteria are as follows:

1. Patients with a chronic foot ulcer of at least 6 weeks duration due to leprosy neuropathy.
2. ≥18 years of age.
3. Ulcer surface area between 2 and 20 cm2 inclusive.
4. Ulcer is clean, dry, and free from infection.
5. Patient can provide informed consent.

The exclusion criteria are as follows:

1. Any significant medical condition, laboratory abnormality, or psychiatric illness that would prevent the participants from participating in the study.
2. Ulcer with surface area <2cm2 and >20cm2.
3. Patient requires skin graft.
4. Any condition that confounds the ability to interpret data from the study (i.e., HIV, chronic Hep B, chronic Hep C or TB patients under active treatment).
5. Any wound that has clinical microbial infections.
6. Diabetes or diabetic ulcer
7. A patient has returned to the hospital having already taken part in the trial.
8. The wound dressing protocol for HELP study is attached as a supplementary material.

In the uncommon scenario where a participant has more than one foot ulcer the largest ulcer will be selected as the index ulcer before randomisation, and all ulcers will receive the same treatment (for example, if the participant is in the intervention group all ulcers – not just the one being monitored for the trial – will be treated with honey). Observations will be made on all ulcers but only the largest ulcer will be used in the primary analysis (please see discussion). Eligible patients will be offered entry in the trial at the point where their senior clinician judges them to be suitable for the honey treatment.

This point arises once the lesion has been cleared of any debris or infection, typically by 7-10 days after beginning treatment with or without antibiotic and/or debridement.

Swabs are taken as a routine, but the interpretation of those swabs is under the control of the clinician. We shall record the result of the (trial) swab taken before randomisation.

### Who will take informed consent? {26a}

Researchers at the study centre have been trained on Good Clinical Practice (GCP). They will be responsible for the screening of eligible participants and taking of consent. The participants’ information sheet has been interpreted into the local language (Hausa). All relevant information for the research participants on the study are contained in the participants’ information sheet. The information sheet will be given to the participants a day before enrolment into the study. The content of the information sheet will be explained to them verbally, and they will be encouraged to ask questions should they need more clarification. Written informed consent will be sought from the eligible participants after the study has been explained to them and they have taken time to decide to enrol into the study. The consent form will be signed or thumb/fingerprinted by the participants before enrolment.

### Additional consent provisions for collection and use of participant data and biological specimens {26b}

The patient information sheet contains all information about the data to be collected and how it will be stored and used. Biological specimens will not be collected as part of this trial. Participants who refuse to give consent will continue to receive normal care from the hospital, but they will not be part of the study.

Participants can decide to withdraw their consent at any time during the study. Such participants will be given a form to complete, stating their reason for withdrawal. A participant may either withdraw fully or they may withdraw from treatment but consent to ongoing data collection.

Once a person has been consented to participate in the trial, baseline data will be collected. This will precede randomisation.

Data will be collected directly onto electronic tablets and the computer program will range check information. Photographs of ulcer will be captured using the tablet device. Each participant will be allocated a unique trial number which will be included in all data entry forms and linked to photographs. The level of activity (step count) will be recorded at each dressing change across both intervention and control groups until 84 days or discharge, whichever comes first. This date will enable us to check for post-randomisation (‘performance’) bias.

## Interventions

### Explanation for the choice of comparators {6b}

The participants in the control group will receive usual care of twice-weekly normal saline dressings only. The normal saline dressing is the standard ulcer dressing method currently in use in the hospital where the trial is taking place.

### Intervention description {11a}

The intervention for this study is raw, unadulterated honey obtained from local bee farmers in North-Central Nigeria. The honey samples have been examined at The National Institute for Pharmaceutical Research and Development (NIPRD), Abuja and confirmed to be free from Microbial contaminants. The honey is stored in airtight plastic containers at room temperature, and away from direct sunlight. The honey will be applied topically to the wound during dressing under strict hygienic conditions using sterilized equipment.

The treatment will be applied at the time of twice weekly changes of dressings by local trained nurses or paramedics. These dressing changes are part of routine care and will thus apply to the intervention and control groups. Dressing changes may take slightly longer for participants in the intervention group but pain is unlikely as the affected limb has a loss of sensation. Participants in both groups have twice weekly dressing changes during their hospital stay until ulcers are healed. Any missed sessions will be noted but this will not be treated as a protocol deviation.

Interval pilot to monitor blinding of observations

We are concerned that honey residue may be discernable by the observers of the ulcer outcome. We will therefore conduct an interval pilot to examine this issue. To ensure proper blinding of the trial, images of the ulcers from the 2^nd^ dressing change (following the honey or saline application at the 1^st^ dressing change) for the first 10 enrolled participants will be sent to 3 independent assessors in Nepal. The assessors will look out for traces of honey residues that might interfere with blind assessment during the study. If the assessors are able to distinguish between cases treated with honey and controls, then we will consider alternating honey and saline dressings and making weekly observations rather than twice weekly observations.

We will also make a video recording of the first dressing change in the first 5 intervention and 2 control participants and send it to the 3 independent assessors. An uninterrupted videorecording will be made from before the dressing is removed until after it has been replaced.

### Criteria for discontinuing or modifying allocated interventions {11b}

None.

### Strategies to improve adherence to interventions {11c}

None.

### Relevant concomitant care permitted or prohibited during the trial {11d}

In line with standard practice, participants will be discouraged from bearing weight on the ulcer site since weight bearing and level of activity of patient might affect the rate of healing of the ulcers. Participants will be asked to wear pedometers on the non-affected foot and the level of activity will be recorded on the tablet at each dressing change from the first dressing change until 84 days from randomisation or discharge, whichever comes first.

### Provisions for post-trial care {30}

The date of discharge will be noted along with participant contact details, address, and contact details for at least one family member. The participants will be routinely contacted six months after randomisation. The treated ulcer area will be examined and photographed. The researcher will take photographs of the ulcer area (healed in the majority of cases). The dates covering any re-admissions to any hospital for treatment of the ‘trial ulcer’ will be recorded.

### Outcomes {12}

We define two main outcomes relating to ulcer healing. The main endpoints will be:

1. Rate of healing based on two observations per week (until healed or 84 days from randomization)
2. Time to healing defined as complete re-epithelization (up to 84 days). Secondary endpoints:
3. Long-term (6 months from randomisation) endpoints will be:
  I. Recurrence of treated ulcer.
  II. Appearance of a new ulcer. Long-term endpoints will be measured at the time of follow up at 6 months from randomisation.
  III. Days hospitalised prior to discharge and total (to include any readmission related to leprosy-ulcers) by 6 months.
  IV. Number of visits to any healthcare facility from discharge to the end of follow-up at 6 months.

### Participant timeline {13}

Participants’ timeline through the trial is outlined in Fig. 1

### Sample size {14}

Recruitment will continue for 24 months. We expect to enroll between 90 and 130 participants. Sample size is based on the two clinical outcomes: rate of healing and time to complete re-epithelialization. We expect at least 70% of ulcers to heal within 84 days with standard care (according to a recent study of neuropathic ulcers, over half due to leprosy [18]). If we assume that the intervention will increase this proportion to 90% and hazards are constant and proportional (so that the hazard ratio is 1.91 for discharge), for a two-sided test of the hazard ratio with a type I error of 5% and statistical power of 80% and a 1:1 allocation ratio, 47 individuals are required in each group. With 130 participants, this allows for a drop out rate of up to 40% to achieve power of 80%. At the most pessimistic sample size of 90, with a drop out rate of 40%, our minimum detectable effect size (i.e. the effect size that achieves an 80% power with 33 patients per arm) is a hazard ratio of 2.15, or 92.5% of patients in the treatment group being discharged by end of the trial period. At the most optimistic sample size of 130 with no drop out, our minimum detectable effect size is a hazard ratio of 1.74, or 87.3% of patients being discharged in the treatment group. All calculations are based on a log-rank test.

We take a conservative approach and base the sample size calculation on the healing outcome and model with lower efficiency to ensure an adequate sample size for all main outcomes since our inferential approach is not based on statistical significance but on a consideration of effect sizes and a comparison and triangulation of the totality of evidence. We are not concerned particularly with being “overpowered” or of a problem of multiple comparisons, but on ensuring a sufficient sample size to estimate clinical effectiveness to a reasonable degree of precision [19;20].

### Recruitment {15}

Eligible individuals are identified by on-site clinical research team and are invited to take part in the study.

### Assignment of interventions: allocation

#### Sequence generation {16a}

Participants will be enrolled sequentially, stratified by ulcer size (above or below 10 cm^2^) and randomly allocated (1:1) to undergo honey treatment or usual care with normal saline using a “digital sealed envelope” method [21].

#### Concealment mechanism {16b}

An allocation table will be generated remotely at the trials office at The University of Ibadan. A permuted block randomisation method will be used to generate the randomization sequence within each stratum. Randomly selecting blocks of size 2, 4, 6, or 8 will be generated in order to maintain balance between the numbers allocated to each of the two groups and to ensure allocation concealment. The generated table will be uploaded into the REDCap database platform to be used for participant enrolment. Access to the allocation table will be restricted. Trial staff in Nigeria will not have access to the allocation table. When a participant’s details are submitted, the trial arm and a unique study number will be assigned and revealed to the local clinician so that the randomised group that the participant is assigned to cannot be altered.

#### Implementation {16c}

A trial statistician at The University of Ibadan will generate the allocation sequence while researchers and clinical staff on-site will enroll and assign participants to respective control or intervention groups.

### Assignment of interventions: Blinding

#### Who will be blinded {17a}

Only the overall research supervisor on site, the database managers in Birmingham and the University of Ibadan trials unit, the clinical staff carrying out dressing changes in the room designated for this purpose and participants themselves will be aware of participants’ randomly assigned group. Ward staff will not be informed. The assessors in Nepal will be blinded from the treatment provided for randomly assigned groups.

#### Procedure for unblinding if needed {17b}

There is no provision for unblinding of trial participant during the trial.

### Data collection and management

#### Plans for assessment and collection of outcomes {18a**}**

Standardised photographs [22] will be taken twice weekly during in-patient stay dressing changes for participants in both intervention and control groups. Any residual honey will be gently removed with a wet swab. Photographs will be taken using the in-built camera in the tablet devices (Samsung Galaxy Tab S7). The photographs will be taken perpendicular to the ulcer. For calibration purposes, a 3cm size clean paper ruler with date and participant’s trial identification number will be placed in the photograph frame above or below the ulcer but at the level of the skin. The photographs will then be uploaded into the REDCap database platform and accessed by database managers at University of Birmingham. The ulcer photos will then be randomised and sent to the blinded assessors in Nepal for measurement of ulcer dimensions. The photograph will be evaluated digitally by the designated observer in Nepal using the PictZar™ Digital Planimetry Software with an electronic PUSH Tool (National Pressure Injury Advisory Panel (NPIAP) at https://npiap.com/page/PUSHTool). The observer will delineate an area of interest by manually ‘painting’ the ulcer area with colour using a computer mouse. The software will then calculate the ulcer dimensions based on this profile.

The assessors will be ‘blinded’ to the treatment allocated to the participant. The assessors are all experienced in leprosy, ulcer care and measurement of ulcers using the PictZar™ software. All photographs from a given participant will be assigned to the observers at random. So that the measurements are not all relegated to the end of the study, they will be made in batches of ten participants reaching completion of their treatments. A proportion (20%) of all ulcer photographs will be measured by two assessors to estimate inter-rater reliability. These photographs will be selected at random.

Treatment will stop when the local clinician determines that complete epithelisation has taken place. Photographs will be taken at this point and at the follow up visit.

#### Plans to promote participant retention and complete follow-up {18b}

The Leprosy Mission Nigeria has a meal subsidy programme for all in-patients at The Leprosy Referral Hospital, Chanchaga. The meal subsidy programme will continue through the period of this study. It is expected that the meal subsidy programme will be sufficient to keep the patient at the hospital during the period of admission up to the time of discharge. Phone contacts of participants or their close relatives will be collected and used for post-discharge follow-up.

#### Data management {19}

All data generated from this study will be classified according to the University of Birmingham Information Security Framework. All data will be collected and stored electronically to reduce data entry errors, such as contradicting answers. Data will be reported on an electronic Case Report Form (eCRF), and all local staff will be trained to collect data directly onto electronic tablets. Data will be acquired and stored on the REDCap database platform with access restricted by passwords at both the University of Ibadan and the local site in Nigeria. Each participant will be allocated a unique study number when they agree to participate (and before randomisation), which will be used on all documents. A master list linking a trial participant number to their identity (name) will be retained by the hospital securely in a locked filing cabinet. This is necessary so that the notes of any person who withdraws consent for data storage can be removed. The trial participant master list will be stored separately from patient lists and the trial data.

#### Confidentiality {27}

Participant confidentiality will be ensured throughout the study and no subject-identifying information will be released to anyone outside the project. Confidentiality will be secured through several mechanisms. Each participant will be assigned a study subject ID, which will be used on all study forms. Any study forms and paper records that contain personal identifier information (e.g., address lists, phone lists) will be kept secured and locked at trial site. Access to all participant data and information at the sites will be restricted to authorized personnel.

Participants will not be identified by name in any reports or publications. Nor will the data be presented in such a way that the identity of individual participants could be inferred. Analysis files created for further study by the scientific community will have no participant identifiers.

#### Plans for collection, laboratory evaluation and storage of biological specimens for genetic or molecular analysis in this trial/future use {33}

This trial will not collect or retain biological specimen for any future use.

### Statistical methods

#### Statistical methods for primary and secondary outcomes {20a}

Time to healing will be analysed using a Cox proportional hazards model with and without adjustment for baseline characteristics (trial ulcer area and participants’ age) allowing for right-censoring. For the rate of healing, we will define the outcome ulcer size in cm^2^ at each time point and include in the model time since admission, treatment status, and their interaction. We will analyse this model using a linear mixed-effects model with participant-level random effects and both with and without adjustment for participant characteristics. Given there are multiple clinical outcomes (two outcomes, with and without adjustment) we will adjust reported p-values for multiple testing using a stepdown method, which provides an efficient means of controlling the family-wise error rate [23]. We will derive the approximate distributions of the test statistics to perform the stepdown procedure using a permutation test approach, by simulating 10,000 re-randomisations of the individuals [24].

#### Interim analyses {21b}

An interim analysis will be conducted when at least 49 (three eights of the target) have been followed up for of 84 days or discharged, whichever comes first. The rationale for this analysis is the detection of a ‘penicillin like’ benefit or statistically significant negative effect of the treatment on either primary outcome. The statistical methods will be as specified above. A statistical threshold of 0.01, one-sided, (0.02 two-sided) will be used for either primary outcome. If either threshold is exceeded, or if the Independent Data Monitoring Committee (IDMC) has other concerns, then the Trial Steering Committee will be advised accordingly. If the IDMC wishes to meet again before trial conclusion they will be able to do so. Only the trial statistician and the Independent Data Monitoring Committee will see the ‘unblinded’ data, unless the Trial Steering Committee are informed.

#### Methods for additional analyses (e.g. subgroup analyses) {20b}

We will also compare average daily step count between treatment and control groups as a simple difference in means (t-test). Since one group may stay longer in hospital than the other and since there may be an interaction between rate of healing and step count, we will compare step counts over periods pre-set at 7, 14 and 42 days.

#### Methods in analysis to handle protocol non-adherence and any statistical methods to handle missing data {20c}

In the unlikely event of missing data by any reason such as withdrawal of participant before completion of treatment, and that the rate differs between groups, a sensitivity analysis will be carried out. The pattern and rates of missing data that are particularly related to the endpoints will be explored. Strategies such as multiple imputation within the sensitivity analysis will be explored instead of imputing missing values.

#### Plans to give access to the full protocol, participant level-data and statistical code {31c}

The full protocol, non-identifiable participant level data and statistical code may be available for sharing once the trial has ended. All requests will be approved by the Chief Investigator (CI), Professor Richard Lilford.

### Oversight and monitoring

#### Composition of the coordinating centre and trial steering committee {5d}

The Leprosy Mission Nigeria is the study sponsor and will oversee the study process in Nigeria. The University of Ibadan will provide data management services and perform quality checks while the University of Birmingham will house the data securely and perform quality checks.

The Trial Management Group (TMG) includes individuals at the University of Birmingham, The Leprosy Hospital Chanchaga, The Leprosy Mission Nigeria, the German Leprosy and TB Relief Association/RedAid Nigeria and the University of Ibadan Clinical Trials Unit who are responsible for the day-to-day management of the trial. The TMG will meet monthly by teleconference, but this may be more frequent if deemed necessary by the members.

The Trial Steering Committee (TSC) provides overall supervision of the trial and will ensure that it is being conducted in accordance with the principles of Good Clinical Practice and other relevant regulations. The TSC will have oversight of the trial. It will send its report to the overall RIGHT programme steering committee, but the TSC will have the final say with the running of the trial itself. The Committee will meet either face to face or via teleconferencing. Meetings will be scheduled for before enrolment of participants begins and following each meeting of the Independent Data Monitoring Committee and then during the analysis phase or more often if required.

#### Composition of the data monitoring committee, its role and reporting structure {21a}

The Independent Data Monitoring Committee consist of individuals who have no conflict of interest within the study. Safety and efficacy data will be supplied, in strict confidence, for review by the Independent Data Monitoring Committee after the 49^th^ participant has have been followed up for of 84 days or discharged, whichever comes first. The Committee will be asked to give advice on whether the accumulated data from the trial, together with the results from other relevant research, justifies the continuing recruitment of further participants. The Committee will meet either face-to-face or by teleconferencing. An emergency may also be convened if a safety issue is identified. The IDMC will escalate any issues and recommendations to the Trial Steering Committee who will make decisions based on these recommendations.

#### Adverse event reporting and harms {22}

The principal investigator in Nigeria will be responsible for recording all Adverse Events (AEs) and reporting any Serious Adverse Events (SAEs) to the University of Birmingham and the University of Ibadan within 24 hours of the research staff becoming aware of the event. An SAE Form will be available on the tablets used to collect data and we will maintain a database of all safety/adverse events. The forms will be reviewed by the TMG which meets monthly and if required, also by the Project Manager. The TSG will periodically review all safety data and liaise with the Independent Data Monitoring Committee regarding any safety issues.

Any deaths will be reported to the Sponsor irrespective of whether the death is related to the disease progression, the intervention or an unrelated event. Only deaths that could plausibly be caused by the intervention will be reported to the Sponsor immediately.

#### Frequency and plans for auditing trial conduct {23}

The trial is audited and monitored by the Sponsor, TLM Nigeria.

#### Plans for communicating important protocol amendments to relevant parties (e.g. trial participants, ethical committees) {25}

Any protocol amendment will be reported to the TMG to approve the change. The amendment will be sent to the sponsor to confirm substantiality and then to National Health Research Ethics Committee (NHREC) for approval.

### Dissemination plans {31a}

The results of the trial will first be reported to trial collaborators. The main report will be drafted by the trial coordinating team and the final version will be agreed by the Trial Steering Committee before submission for publication, on behalf of the collaboration.

We will present our work at conferences such as the annual conference of the Neglected Tropical Disease, NGDO Network (NNN) of which The Leprosy Mission is a member, Health Systems Global Conference and the International Leprosy Congress in 2024. Other dissemination plans include bite-sized research reports in lay format, public announcements in communities in LMICs, policy briefings, print and online media, the Chief Investigator’s News Blog (680+ subscribers), institutional and professional social media accounts and websites.

## Discussion

This study protocol describes a clinical trial with honey as intervention for dressing of ulcers in leprosy. It is a single centre, comparative, prospective, single blinded, parallel group, blocked stratified randomised controlled trial.

Leprosy is regarded as a Neglected Tropical Disease (NTD) because of its near absence from global health agenda [25], as such, very little attention has been given to treatment of ulcers in leprosy in spite of its debilitating nature and the social stigma it attracts. Methods such as laser therapy [26], zinc tape [27], pentoxifylline injection [28], amniotic membrane gel [29], phenytoin suspension [30], and Leukocyte-platelet rich fibrin [3] have been tested on leprosy foot ulcers as the search for the most suitable treatment continues. Normal saline has been the most common comparator in most of the studies on ulcers in leprosy [31]. Amongst the challenge with the normal saline dressing are the slow rate of healing, the possibility of ulcers getting infected with pathogens [14], and the likelihood of recurrence of treated ulcers. A Cochrane review [31] noted the limitation of most previously published evidences to be high or unclear risk of bias (due to selection, performance, detection, or attrition), imprecision due to few participants in the study, indirectness from poor outcome measurement, and inapplicable interventions.

Although honey is known for centuries to promote wound healing, there are only few literatures that discuss its usage in controlled clinical trials. While honey is noted to have wound healing properties including debridement, deodorisation, and antimicrobial properties [15], its usage may result in painful sensation in about 5% of persons, although this is unlikely to occur in leprosy and diabetes [16]. The hypothetical risk of botulism will be mitigated by regularly screening the honey sample. Detailed procedures for the study have been described in this protocol document. We expect that the outcome of this study will provide valuable evidence on the use of honey for the treatment of ulcers in leprosy.

## Data Availability

All data produced in the present study are available upon reasonable request to the authors

## Trial status

Protocol version number: 0.9; 2 December 2022.

Recruitment of trial participants begins on 14 March 2022. The recruitment will continue for approximately 24 months.

## Abbreviations

AEs: Adverse Events
CI: Chief Investigator
GCP: Good Clinical Practice
HELP: Honey Experiment on LeProsy Ulcer
IDMC: Independent Data Monitoring Committee
MDT: Multidrug Therapy
NGDO: Non-governmental Development Organisation
NHREC: National Health Research Ethics Committee
NIHR: National Institute for Health Research
NIPRD: National Institute for Pharmaceutical Research and Development
NNN: Neglected Tropical Disease NGDO Network
NTD: Neglected Tropical Disease
PI: Principal Investigator
RIGHT: Research and Innovation for Global Health Transformation
SAEs: Serious Adverse Events
TLM: The Leprosy Mission
TLMN: The Leprosy Mission Nigeria
TMG: Trial Management Group
TSC: Trial Steering Committee

## Declarations

## Acknowledgements

We acknowledge the contribution of the public health department of Niger state ministry of health and the Chanchaga Leprosy referral hospital to the clinical trials. We also acknowledge the members of leprosy affected communities in Nigeria for the assurance of cooperation during pre-trial engagements. We thank the Chair and members of the trial steering committee: Dr Doyin Odubanjo, Professor Kara Hanson, Dr Paul Saunderson, Dr Jerry Joshua for their constructive advice on the trial. We also acknowledge the support of Dr Indra Napit, Dilip Shrestha, Karuna Neupane and Anjali Shrivastav from The Leprosy Mission Nepal for their support in training for the trial.

## Authors’ contributions {31b}

POS, PAT, IOO, AM, and LU are local co-investigators and have contributed to the study conception, design, and intellectual content of the protocol. SC is the research project manager at the University of Birmingham and has contributed to the study conception, design, and intellectual contents of the protocol. JS is the study senior project manager at the University of Birmingham and has contributed to the study conception, design, and intellectual contents of the protocol. OI is research fellow in the study and has contributed to the study design, and intellectual contents of the protocol. SIW contributed to the design of the evaluation, sample size calculation, randomisation and quantitative sample selection, and critically evaluated the intellectual content of the protocol. MA is the project manager at the University of Ibadan, and critically evaluated the intellectual content of the protocol. JA contributed to the design of the trial randomisation and intellectual content of the protocol. AO leads the clinical trials team at the University of Ibadan and contributed to the study design and intellectual content of the protocol. SOU is the local chief investigator and contributed to the study conception and intellectual content of the protocol. RJL is the principal investigator at the University of Birmingham and contributed to the study conception and design, and critically evaluated the intellectual content of the protocol. All authors read and approved the final manuscript.

## Ethics approval and consent to participate {24}

The trial will be conducted in full conformance with the principles of the Declaration of Helsinki and Good Clinical Practice (GCP) Guidelines. It will also comply with all applicable UK legislation and Standard Operating Procedures for University of Birmingham Sponsored Studies. Ethics approval has been granted in Nigeria for the study by the National Health Research Ethics Committee (NHREC) (Approval number: FHREC/2022/01/09/04-02-22), and Niger State Ministry of Health (Reference number: STA/495/Vol/199).

## Consent for publication {32}

No identifying information on any individual’s data will be presented. All participants will have the opportunity to review information about the study and give informed consent. A model consent form will be provided on request

