## Supplementary material for "Honey Experiment on LeProsy Ulcer (HELP) Study Protocol: A Randomised Control Trial of Raw, Unadulterated African Honey for Ulcer Healing in Leprosy: A Study Protocol"

### Appendix 1

**Wound dressing protocol for HELP study at Leprosy Referral Hospital Chanchaga**

If Patients fulfil the inclusion criteria and offer to enrol them in the study.

**Screening and consent taking:-**

1.	Participant admitted to hospital.

2.	Check the Patients’ vitals. Note down all parameters.

3.	Send ulcer swab for culture to lab and check for any microbial growth and their antibiotic sensitivity. Irrigate wound with normal saline before taking swab.

4.	After receiving the reports, screen the participants for eligibility using REDCap and record size of ulcer- length, breadth, depth and area in cm/cm^2^ using measurement scale.

5.	If participant is eligible and willing to take part in the trial, take informed consent.

6.	The patients should have the details of the procedure explained to them on the first day and written consent should be provided the following day with signature or fingerprints.

7.	Collect baseline data, following consent, before randomization

**Wound Preparation for both Control and Intervention Groups:**

1.	Fill out the participant e-CRF (case report form) into REDCap.

2.	Record time as hour: minutes so that we can calculate the time taken for dressing change. It should be the duration of time from opening the dressing until closing the wound with bandaging.

3.	Clean the wound with normal saline through irrigation to remove all exudates.

4.	Wound debridement - remove the devitalized tissue and fibrin membrane surrounding the ulcer by careful debridement (marginal resurfacing) of the wound. Clean the wound with normal saline. Ulcer debridement is not recommended at every dressing change.

5.	Take photographs perpendicular to wound with a labelled (Participant Identification Number and Date) 3cm ruler at the level of the skin and calculate measurements using PUSH tool, and record these locally. Record the wound- length, breadth, depth and surface area in cm/cm^2^. Measurements should not be recorded in REDCap software by the Nigeria team, as wound assessment will be performed by a team from TLM Nepal blind to intervention status. Where an index ulcer heals into multiple smaller ulcers, the surface area of all ulcers within the original index ulcer site will be recorded in cm^2^.

**Randomization:**

All participants will be allocated into 2 groups according to computer generated randomisation schedule:

1.	Twice a week dressing with normal saline (Control group)

2.	Twice a week dressing with Honey (Intervention Group)

**Honey Application to the Wound for Intervention Group:**

1.	Filter the honey into sterile container using sterile polypropylene 0.5 micron filter cloth.

2.	Prepare clean dry gauze and soak with the filtered honey. Apply the honey-soaked gauze directly on the wound.

3.	Cover the wound with sterile Vaseline gauze and apply sterile dry gauze on it, close wound with bandaging. Apply liquid paraffin to the surroundings of the wound if the foot skin is dry. Convince the participants to avoid weight bearing at the wound site. There is no need of dressing every day. Record the total time taken for dressing change as hour: minutes.

4.	Open the wound after 3-4 days as per twice a week dressing schedule. Clean the wound with normal saline, dry it with gauge, and do not rub the wound. Take photographs, note down the measurement of wound size and depth.

**Normal Saline Wound dressing for Control Group:**

Wound preparation procedure is same as mentioned above.

1.	Perform irrigation with 50-100 ml Normal saline depending on the size of the ulcer. Apply normal saline gauge over the wound area and cover with sterile Vaseline gauze. Apply sterile dry gauze over it and close wound with bandaging. Apply liquid paraffin to the surroundings of the wound if the foot skin is dry. Convince the participants to avoid weight bearing at the wound site. There is no need of dressing every day. Record the total time taken for dressing change as hour: minutes.

2.	Open the wound after 3-4 days as per twice a week dressing schedule. Repeat the Normal saline dressing as mentioned above. Do not rub the wound surface. Take photographs, note down the measurement of wound size and depth.

**Implement following to all participants in both Intervention and Control groups-**

Provide a glass of energy drink to participants every time after dressing change.

Add Iron tablets (200 mg BD), folic acid (5 mg OD), Vitamin C (500 mg BD) and Multivitamins (1 tab BD) to the participant’s medication list during admission period.

**Post wound healing care and Follow up:**

1.	If the wound is completely epithelialized before 70 days after enrolment, check for the consistency of tissue at wound site. Take a photo with PUSH tool tablet camera of whole foot. Apply Vaseline gauge at healed wound site and liquid paraffin around it, then apply Plaster of Paris (PoP) cast with window at healed wound site for 2-4 weeks until the healed ulcer site will have matured tissue.

Participants will be discharged from hospital after POP application. They can be transferred to Self-Care Unit until the PoP will be removed out or can be sent to their home. The POP should be removed after 2-4 weeks.

2.	After removal of PoP, train the participants for self-care and prevention of disability.

3.	Consult footwear staff to design protective footwear.

4.	Discharge the participants from Self-Care Unit and ask them to come at six months from randomisation for follow up evaluation.

5.	At follow up check condition of foot and record any recurrence or new ulcer at same foot.

6.	Take photographs with PUSH tool at follow-ups.

7.	If any complications or adverse events are seen during admission period, please inform the Principal Investigator or RIGHT Research Staff.

### Appendix 2

**Honey Experiment on LeProsy Ulcer (HELP): A Randomised Control Trial of Raw, Unadulterated African Honey for Ulcer Healing in Leprosy**

**Participant Information Sheet**

**Introduction**

We would like to invite you to take part in a research study. Joining the study is entirely up to you. Before you decide, you need to understand why the research is being done and what it would involve. One member of our team will go through this information sheet with you, and answer any questions you may have. Ask questions if anything you read is not clear or you would like more information. Please feel free to talk to others about the study if you wish. Take time to decide whether or not to take part.

**Who is organising and funding the study?**

The study is being organised by The Leprosy Mission Nigeria in collaboration with the German Leprosy and TB Relief Association and the University of Birmingham, UK. The study is funded by the UK National Institute for Health Research.

**What is the purpose of the study?**

Leprosy ulcers are not caused by the leprosy germ but by loss of sensation leading to repetitive injury. Treatment consists of keeping the ulcer clean and fresh and also applying wet bandages regularly – dressing changes. You will currently have these dressing changes every 3 or 4 days.

The purpose of our study is to test a new method that may help the ulcer to heal faster. This treatment is done while you have your dressing changed. We will use honey to dress the wound for some of you while others will receive normal saline dressing on their wounds. The selection of participants for the honey or normal saline dressing will be done randomly. Honey is being used for wound dressing for centuries but this time, we want to do it as a registered trial.

At present, this treatment appears to be very safe, although we do not know if it works. This study only looks at ulcers on the feet or legs and not anywhere else.

**Why have I been asked to take part?**

You have been invited because you have ulcer in your foot.

**Do I have to take part?**

No. It is up to you to decide to take part or not. If you don’t want to take part, that’s ok. Your doctor will still care for you and your decision will not affect the quality of care you receive.

We will discuss the study together and give you a copy of this information sheet. If you agree to take part, we will then ask you to sign a consent form.

**What will happen to me if I take part?**

If you are willing to take part in this study, we will first ask you to sign a consent form which is your indication that you understand the study and agree to take part.

Then you will be evaluated, do an interview and physical examination of your ulcer in your foot. If you meet the study’s criteria, and you wish to participate, you will receive dressing with normal saline or with honey. The treatment will be chosen by chance by a computer so that half of the people in the trial get the normal saline (control group) and the other half get the honey (intervention group). It is really important that the two groups for this study have a similar mix of patients in them. Having a similar mix means that we know that if one group of participants does better than the other, it is very likely to be because of the treatment and not because there are differences in the types of patients in each group. You will have an equal chance of receiving either normal saline dressing or honey dressing.

It is important that you realise that treatment is not always effective. If you agree to take part of this study, we will ask you to complete different questionnaires. You will be called for follow-up six months after randomisation for the trial.

**What will I have to do?**

You will be expected to be admitted in hospital during the treatment period. You have to answer the entire questions asked to you. This will help us to gather information about you and your progress during the study period.

**What information will be collected?**

Only simple information about you, your treatment for your ulcer and how it affects you will be collected. This will include your name, but you will only ever be viewed by your participant number. We will keep this information separate from your address.

We will also take photographs of your ulcer every time you have your dressing changed to see how well it is healing. These photographs will only ever be viewed by your participant number. We may also make video of your dressing change.

**What will happen to information collected about me?**

All information collected about you will be kept private. Only the study staff and authorities who check that the study is being carried out properly will be allowed to look at information about you. Data may be sent to other study staff at University of Birmingham but this will be anonymised. This means that any information about you which leaves the hospital/surgery/clinic will have your name and address removed so that you cannot be recognised.

Your doctor will send some details about you to the study team at university of Birmingham, who will store it securely. Your personal details will be kept in a different safe place to the other study information and will be kept for at least 10 years after study completion. All the data will be securely stored in safe place.

The collected data may also be used for future research, including impact activities following review and approval by an independent Research Ethics Committee and subject to your consent at the outset of this research project.

For further information, please refer to the University of Birmingham Research Privacy Notice which is available here: <https://www.birmingham.ac.uk/privacy/index.aspx> or by contacting the Information and Data Compliance Team at:.

**What if something goes wrong?**

If you have a concern about any aspect of this study, you should ask to speak to the researchers who will do their best to answer your questions. You can also contact Dr. Sunday Udo who is the principal investigator of this study for any queries. If you remain unhappy and wish to complain formally, you can do this by contacting Professor Richard Lilford, University of Birmingham UK,

The study holds insurance policies which apply to this study. If you experience harm or injury as a result of taking part in this study, you may be eligible to claim compensation.

**Can I change my mind about taking part?**

Yes. You can withdraw from the study at any time. You just need to tell your doctor that you don’t want to be in the study anymore. Your doctor will still care for you.

You can withdraw from treatment but keep in contact with us to let us know your progress. Information collected may still be used.

This would not affect the care you receive. If the intervention proved effective, you will be eligible to receive it if you develop a new ulcer or if your ulcer has not healed or recur.

**What will happen to the results of this study?**

The study results will be published in a medical journal so that other doctors can learn from them. Your personal information will not be included in the study report and there is no way that you can be identified from it.

**Who has reviewed the study?**

All research involving human participants is looked at by an independent group of people, called a Research Ethics Committee, to protect your interests. This study has been reviewed and given favourable opinion by University of Birmingham’s Science, Technology, Engineering and Mathematics (STEM) ethics committee.

**Who should I contact if I want further information?**

Professor Richard Lilford, University of Birmingham, UK

*****Thank you for taking time to read this information leaflet. If you think you will take part in the study please read and sign the consent form.*****

### Appendix 3

**CONSENT FORM**

**Title of Project:** **Honey Experiment on LeProsy Ulcer (HELP): A Randomised Control Trial of Raw, Unadulterated African Honey for Ulcer Healing in Leprosy**

**Name of Researcher(s):** Dr Sunday Udo, TLM Nigeria and Professor Richard Lilford, University of Birminigham, UK

A.	I __________________________________________________________________ understand that doctors at the Leprosy Referral Hospital Chanchaga/St Benedict’s TBL and Rehabilitation Hospital Ogoja, and researchers at University of Birmingham, University of Ibadan, The German Leprosy and TB Relief Association, and The Leprosy Mission Nigeria and Nepal are involved in research into alternative treatments methods for leprosy ulcers. This study will look at whether honey is more beneficial than normal saline dressing in healing foot ulcer or not. We are hoping to prove efficacy of new treatment method for healing leprosy ulcer.

B.	The study has been explained to me.

C.	I confirm that I am 18 years old or above.

D.	I shall be randomly assigned to a normal saline dressing group or honey dressing group. There is equal chance of getting either normal saline dressing or honey dressing.

E.	I agree to have photographs and videos taken during the ulcer dressing.

F.	I agree that my collected data be used for further research in future*.

*Please note that participants may say ‘NO’ to this question and still take part in the study.

G.	I can decide to leave the study at any time for any reason and will still receive other treatment from the hospital for my condition.

H.	I understand that my name will not be revealed in any published material concerning this study. I understand that my notes will be treated with maximum confidentiality and will only be accessed by staff directly involved in the Study or the monitors of the Study.

I.	I have received enough information about the study in a language I understand. I had the opportunity to discuss it and ask questions, and my questions have been answered to my satisfaction. I understand that participation is voluntary and that I am free to withdraw my consent at any time. I freely consent to participate in this research study and to allow treatment and tests to be performed on me as explained.

J.	I understand that I can be requested anytime to terminate my participation in the trial if the need arises. I will be given full explanation of the reason and will still receive standard treatment.

K.	I agree to take part in the study.

Printed Name	& Signature (or finger print)	Date

Name of Participant ________________________________

Signature/Finger Print	____________________________	_____/______/20____

Name of Witness ________________________________

Signature/ Finger Print ____________________________	_____/______/20____

Name of Researcher______________________________

Signature	________________________________	_____/______/20____
